# Retrospective *in silico* HLA predictions from COVID-19 patients reveal alleles associated with disease prognosis

**DOI:** 10.1101/2020.10.27.20220863

**Authors:** René L. Warren, Inanç Birol

## Abstract

**Background:** The Human Leukocyte Antigen (HLA) gene locus plays a fundamental role in human immunity, and it is established that certain HLA alleles are disease determinants.

**Methods:** By combining the predictive power of multiple *in silico* HLA predictors, we have previously identified prevalent HLA class I and class II alleles, including DPA1*02:02, in two small cohorts at the COVID-19 pandemic onset. Since then, newer and larger patient cohorts with controls and associated demographic and clinical data have been deposited in public repositories. Here, we report on HLA-I and HLA-II alleles, along with their associated risk significance in one such cohort of 126 patients, including COVID-19 positive (n=100) and negative patients (n=26).

**Results:** We recapitulate an enrichment of DPA1*02:02 in the COVID-19 positive cohort (29%) when compared to the COVID-negative control group (Fisher’s exact test [FET] p=0.0174). Having this allele, however, does not appear to put this cohort’s patients at an increased risk of hospitalization. Inspection of COVID-19 disease severity outcomes reveal nominally significant risk associations with A*11:01 (FET p=0.0078), C*04:01 (FET p=0.0087) and DQA1*01:02 (FET p=0.0121).

**Conclusions:** While enrichment of these alleles falls below statistical significance after Bonferroni correction, COVID-19 patients with the latter three alleles tend to fare worse overall. This is especially evident for patients with C*04:01, where disease prognosis measured by mechanical ventilation-free days was statistically significant after multiple hypothesis correction (Bonferroni p = 0.0023), and may hold potential clinical value.

## INTRODUCTION

Modern history has been plagued by deadly outbreaks, from the recurring influenza (e.g. Spanish, Asian, Hong Kong, Avian) and HIV/AIDS viral pandemics, to bacterial and protist infections causing tuberculosis and malaria. Since the early 2000s, we have faced another threat: novel coronavirus infections causing severe respiratory illnesses such as SARS, MERS and today, coronavirus disease 2019 − COVID-19 [1]. The SARS-CoV-2 coronavirus responsible for the COVID-19 respiratory disease is of particular concern; not only does SARS-CoV-2 spread quickly, the symptoms of its infection, when exhibited, are very similar to that of the cold and flu making it difficult to diagnose, trace and contain. Further, infected individuals are affected differently. For instance, older men (≥65 years old) with pre-existing medical conditions, such as diabetes, appear at increased risk of progressing into the more severe phase of the disease, yet SARS-CoV-2 infections affect all other age groups evenly except occasionally in children and adolescents [2]. Most peculiar is that a high proportion of individuals who tested positive for SARS-CoV-2 are asymptomatic – as high as 43% recorded in Iceland, a rate that appears to vary depending on jurisdictions and populations [3-5]. As of now, the disparity in patient response to SARS-CoV-2 infection is still eluding us.

The most efficient way to combat pathogens has been through the use of our own defense mechanism: our acquired immunity. This is done by vaccination campaigns that effectively prime our immune systems at the population level before we even encounter pathogens. But design of effective vaccines must consider interactions with host immune genes. The Human Leukocyte Antigens (HLA) are a group of such genes encoding surface receptors that bind short peptide epitopes derived from endogenous (class I) or exogenous (class II) antigens, including viral antigens, and they facilitate killer or helper T cells to set off an appropriate immune response. The magnitude of this response varies between patients as populations and individuals have different composition of HLA genes and variable T cell repertoires. As such, HLA induces a bias, which is responsible for documented host susceptibility to disease [6]. Some of the notable associations between HLA and disease are observed in AIDS patients, with certain HLA alleles conferring protection [7]. In other cases, HLA has been implicated with autoimmune diseases and diabetes [8-11]. The exact underlying mechanisms behind these associations are unclear, but there is mounting evidence that bacterial and viral infection may be the trigger for some [10] and that HLA plays a critical role in the viral infection cycle, including viral entry into host cells [12].

Since the beginning of the pandemic, worldwide reports have emerged on HLA associations with COVID-19, including our own [13-19]. Using publicly available metatranscriptomic sequencing data made available at the pandemic onset, we had demonstrated the utility of a high throughput *in silico* method for characterizing the HLA types of COVID-19 patients from bronchoalveolar lavage fluid and blood samples and reported on prevalent alleles, including the DPA1*02:02P - DPB1*05:01P HLA-II haplotype observed in 7 out of the 8 of patients from two small cohorts. Here, using public RNA-seq sequencing data from a larger COVID-19 patient cohort with clinical outcomes and demographics data [20], we report on HLA alleles with potential diagnostic (DPA1*02:02) and prognostic (A*11:01, C*04:01 and DQA1*01:02) value in 126 hospitalized patients with (n=100) and without (n=26) COVID-19 and present our findings in light of available demographic characteristics using hospitalization and disease severity metrics.

## METHODS

We downloaded Illumina NOVASEQ-6000 paired-end (50 bp) RNA-Seq reads from libraries prepared from the blood samples of 126 hospitalized patients, with (n=100) or without COVID-19 (n=26) (ENA project: PRJNA660067, accessions: SRX9033799-SRX9033924). This data is part of a large-scale multi omics study from the Department of Molecular and Cellular Physiology, Albany Medical College, Albany, NY, USA, with aims to analyze COVID-19 Severity and clinical as well as demographics data was made available by the study authors [20] (GEO accession GSE157103). On each patient RNA-Seq dataset, we ran HLA prediction software OptiType [21] (v1.3.4), seq2HLA [22] (v2.3), and HLAminer [23] (v1.4 targeted assembly mode with defaults) as described [19]. We tallied HLA class I (HLA-I) and class II (HLA-II, supported by Seq2HLA and HLAminer only) allele predictions and for each patient we report the most likely HLA allele (4-digit resolution), indicating HLA predictor tool support (**Additional file 1, tables S1 and S2**).

Looking at class I and II alleles predicted in 10% or more of COVID-19 positive patients (class I, n=17; class II, n=11) we calculated Fisher’s Exact Test (FET), first testing for enrichment in COVID-19 positive vs. negative patients (R function fisher.test, alternative = “greater”). For those same alleles (found in ≥10% patients) and inspecting only the COVID-19 positive cohort, we tested for the probability of patient hospitalization, as measured by the Intensive Care Unit (ICU) metric reported by the original study authors, using FET. We looked further into the risk of hospitalization in COVID-19 patients with vs. without these alleles using the Kaplan-Meier (KM) estimator (R library survival), plotting the probability of remission using the “hospital-free days post 45 day followup (days)” (HFD-45) metric reported by the study author as a proxy for disease severity, with lower HFD-45 numbers indicating worse outcomes. Similarly, we ran the KM estimator using another metric of disease severity, “ventilator-free days”, which captures the most severe cases with COVID-19 patients suffering respiratory deterioration and requiring mechanical ventilation. On each set we calculated the log-rank p-value (R library survminer) and corrected for multiple hypothesis testing (Bonferroni correction) using the number and patient abundance rank of class I (n=17) or class II (n=11) HLA alleles observed in 10% or more of COVID-19 patients. We also inspected the combined influence of HLA alleles and patient demographics data (age, sex, ethnicity) on the hospitalization (ICU negative vs. positive) outcomes of COVID-19 patients, using odds-ratio calculations (R function fisher.test, and applying Haldane correction [24] on zero values, when necessary).

## RESULTS

We collated the HLA class I and class II predictions of three *in silico* HLA predictors derived from the RNA-seq samples of a recent [20] COVID-19 positive patient cohort (n=100) with control patients (n=26) who tested negative for COVID-19 (**Additional file 1, Tables S1 and S2**). Due to the limiting short read length (paired 50 bp) we chose to first report on OptiType [21] and seq2HLA [22] class I and class II predictions, and count the additional allele support from seq2HLA and HLAminer [23]. In all, we identify 17 and 11 HLA class I and class II alleles predicted in 10% or more of COVID-19 patients, respectively (**Tables 1 and 2**). There were many more alleles predicted (133 class I and 101 class II in the COVID-19 positive cohort), but too few patients are represented at lower cut-offs to compute meaningful statistics. First, we looked at the statistical enrichment (Fisher’s Exact Test - FET) of each allele in the COVID-19 positive set, compared to the COVID-19 negative control group. We find HLA-I A*30:02 and HLA-II DPA1*02:02 allele enrichments nominally significant (FET p = 0.0417 and p = 0.0174) at the 5% level (**Tables 1 and 2**). However, when Bonferroni correction is applied for the number of HLA class I allele tests or when the abundance rank is factored in for A*30:02, the test is not significant (n=17, Bonferroni p = 0.7080; n=10, Bonferroni p = 0.4165, respectively). For HLA-II DPA1*02:02, Bonferroni correction finds the test insignificant at the α=0.05 level for the number of hypothesis, but significant for the allele abundance rank (n=11, Bonferroni p = 0.1916; n=2, Bonferroni p = 0.0348).

**Table 1.** HLA-I alleles identified in 10% or more COVID-19 positive patients and statistical tests of enrichment in the Overmyer *et al*. [20] COVID-19 positive (vs. negative) cohort and association with hospitalization. Red font indicates significant associations (Fisher’s Exact Test) not corrected for multiple hypothesis tests.

| HLA-I | COVID+<br>patients | ICU-<br>patients | ICU+<br>patients | COVID-<br>patients | H1:<br>Enriched in<br>COVID+<br>Fisher's<br>Exact Test<br>p-value | H1:<br>Increased risk of<br>hospitalization<br>Fisher's Exact<br>Test p-value |
| --- | --- | --- | --- | --- | --- | --- |
| A*02:01 | 30 | 15 | 15 | 12 | 0.9614 | 0.5862 |
| A*24:02 | 23 | 12 | 11 | 6 | 0.6161 | 0.5000 |
| C*07:02 | 22 | 9 | 13 | 7 | 0.7890 | 0.8865 |
| C*07:01 | 18 | 6 | 12 | 7 | 0.8991 | 0.9668 |
| C*04:01 | 18 | 4 | 14 | 3 | 0.3231 | 0.0087 |
| C*06:02 | 16 | 8 | 8 | 6 | 0.8708 | 0.6071 |
| B*51:01 | 16 | 6 | 10 | 2 | 0.2292 | 0.9143 |
| C*03:04 | 14 | 4 | 10 | 3 | 0.5171 | 0.9796 |
| C*15:02 | 14 | 6 | 8 | 1 | 0.1363 | 0.8060 |
| A*01:01 | 13 | 5 | 8 | 7 | 0.9742 | 0.8832 |
| A*03:01 | 13 | 7 | 6 | 5 | 0.8680 | 0.5000 |
| A*30:02 | 13 | 7 | 6 | 0 | 0.0417 | 0.5000 |
| B*07:02 | 12 | 5 | 7 | 5 | 0.8969 | 0.8217 |
| A*30:01 | 12 | 8 | 4 | 1 | 0.2016 | 0.1783 |
| B*44:02 | 11 | 4 | 7 | 4 | 0.8320 | 0.9001 |
| A*68:01 | 11 | 5 | 6 | 0 | 0.0697 | 0.7377 |
| A*11:01 | 10 | 1 | 9 | 2 | 0.5320 | 0.0078 |

**Table 2.** HLA-II alleles identified in 10% or more COVID-19 positive patients and statistical tests of enrichment in the Overmyer *et al*. [20] COVID-19 positive (vs. negative) cohort and association with hospitalization. Red font indicates significant associations (Fisher’s Exact Test) not corrected for multiple hypothesis tests.

| HLA-II | COVID+<br>patients | ICU-<br>patients | ICU+<br>patients | COVID-<br>patients | H1:<br>Enriched in<br>COVID+<br>Fisher's Exact<br>Test p-value | H1:<br>Increased risk of<br>hospitalization<br>Fisher's Exact<br>Test p-value |
| --- | --- | --- | --- | --- | --- | --- |
| *DPA1\*01:03* | 46 | 21 | 25 | 17 | 0.9812 | 0.8421 |
| *DQA1\*01:02* | 40 | 26 | 14 | 10 | 0.5363 | 0.0121 |
| *DPB1\*04:01* | 22 | 9 | 13 | 10 | 0.9726 | 0.8865 |
| *DPA1\*02:02* | 29 | 16 | 13 | 2 | 0.0174 | 0.3299 |
| *DPA1\*02:01* | 22 | 11 | 11 | 6 | 0.6578 | 0.5952 |
| *DQA1\*05:02* | 19 | 9 | 10 | 3 | 0.2824 | 0.6945 |
| *DPB1\*01:01* | 19 | 10 | 9 | 3 | 0.2824 | 0.5000 |
| *DQB1\*06:11* | 11 | 6 | 5 | 3 | 0.6811 | 0.5000 |
| *DRB1\*13:01* | 10 | 7 | 3 | 4 | 0.8689 | 0.1589 |
| *DQB1\*06:02* | 10 | 6 | 4 | 3 | 0.7348 | 0.3703 |
| *DRB1\*15:01* | 10 | 4 | 6 | 2 | 0.5320 | 0.8411 |

COVID-19 positive patients could be further stratified into those who were hospitalized and admitted to the Intensive Care Unit (**Tables I and 2**, ICU+), and those who were not (**Tables 1 and 2**, ICU-). When computing FET statistics, we find HLA-I A*11:01 and C*04:01 and HLA-II DQA1*01:02 significant at the α=0.05 level (**Tables 1 and 2**; p = 0.0078, p = 0.0087 and 0.0121, respectively) but none remain significant after Bonferroni correction. The Overmyer study authors [20] reported important disease severity metrics (HFD-45 and days without needing mechanical ventilation), which we used to assess the remission probability of COVID-19 patients having a specific allele using Kaplan-Meier estimation. We find patients of the Overmyer cohort with either A*11:01 (**Figure1a**), C*04:01 (**Figure 1b**) or DQA1*01:02 (**Figure 1c**) to be at a significant increased risk of hospitalization (log-rank p = 0.0099, p = 0.0082 and p = 0.0097). When applying multiple test corrections to account for allele abundance rank, only C*04:01 (n=5, Bonferroni p = 0.0410) and DQA1*01:02 (n=2, Bonferroni p = 0.0194) remained significant at the α=0.05 level. When looking at patients needing mechanical ventilators, a severe outcome in COVID-19 disease progression, we only find patients with C*04:01 to be at a statistically significant increased risk (**Figure 1d**, log-rank p = 0.0019). Multiple hypothesis test correction retains the statistical significance of this allele when factoring both the number of HLA-I alleles tested and C*04:01 abundance rank (n=17, Bonferroni p = 0.0023; n=5, Bonferroni p = 0.0095).

**Figure 1.**
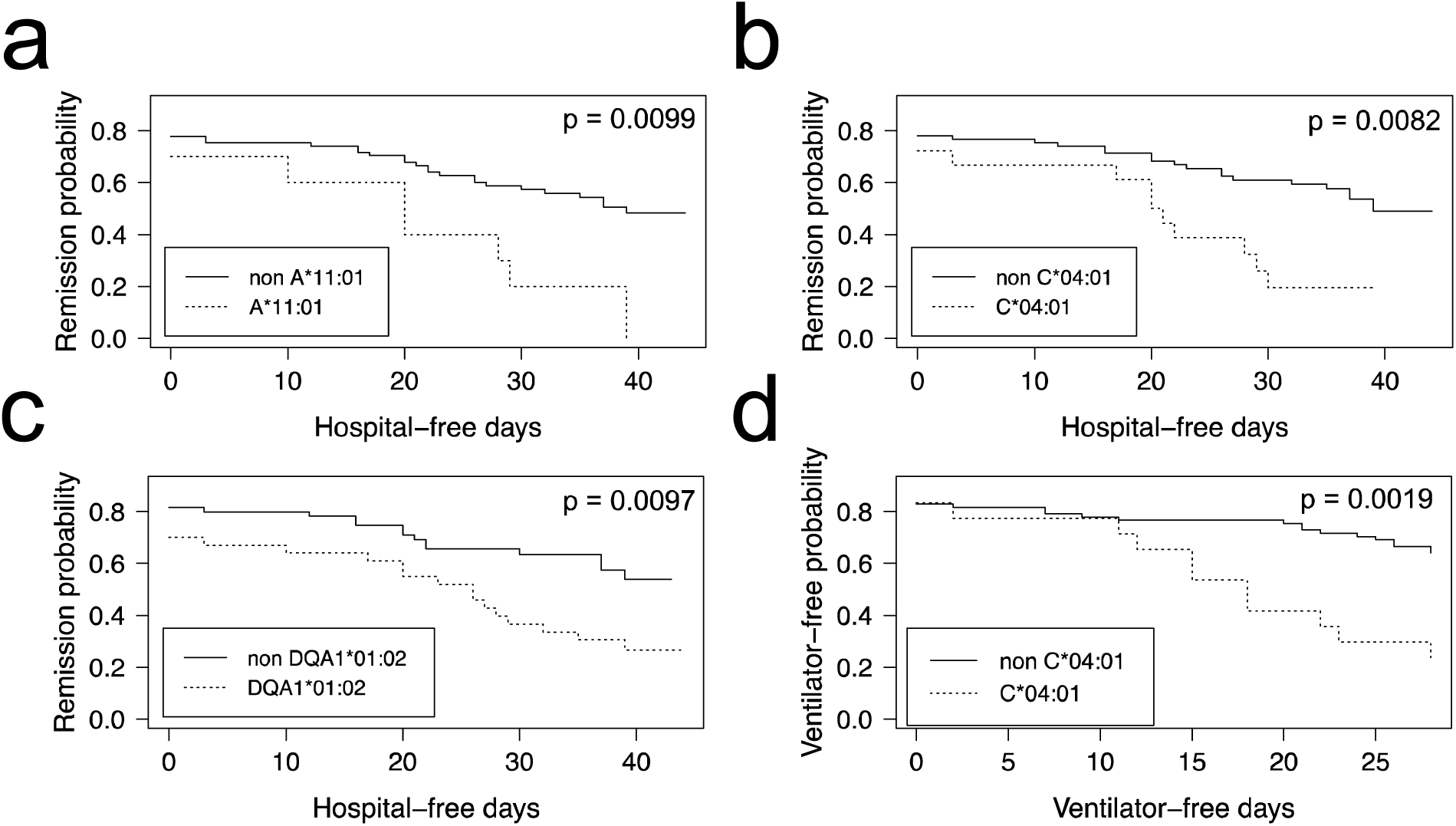
HLA alleles associated with higher risk of hospitalization in a COVID-19 positive patient cohort. COVID-19 positive patients were split into two groups per allele tested, depending on whether they were predicted to have the HLA allele under scrutiny or not. We ran the Kaplan-Meier estimator (R package survival) using the Overmyer *et al*. cohort [20] HFD-45 metric for estimating the remission probability of patients without or with alleles (a) A*11:01, (b) C*04:01 or (c) DQA1*01:02 and mechanical ventilator-free days to estimate the statistical significance of the more severe disease outcome observed in COVID-19 patients with (d) C*04:01. Log-rank p values were calculated for each (R package survminer) and are indicated on the plots.

Looking at the influence of the aforementioned alleles in combination with simple demographics (sex, age and ethnicity), we find that of the Overmyer cohort patients with the DPA1*02:02 allele, those with a white ethnic background and females appear at an increased risk of testing positive for COVID-19 (**Figure 2**; odds ratio [OR] = 6.33 [5.33– 7.34], FET p = 0.0491 and OR = 7.33 [6.18-8.48], FET p = 0.0326, respectively). The association with gender is also observed in alleles A*11:01, C*04:01 and DQA1:01:02, putting female COVID-19 patients of this cohort at an increased risk of hospitalization for the class I alleles (**Figure 2**; OR = 12.09 [10.41-13.76], FET p = 0.0105 for both) and male COVID-19 patients at increased risk for DQA1*02:02 (**Figure 2**; OR = 2.74 [2.69-2.79], FET p = 0.0481). For patients with the latter HLA-II allele, minorities and younger individuals (<65 years old) are also more at risk of hospitalization (**Figure 2**; OR = 4.08 [2.85-5.32], FET p = 0.0222 and OR = 3.62 [2.42-4.83], FET p = 0.0240, respectively). In this cohort, we also find patients with A*11:01 in the younger age group (<65 years old) at increased risk of hospitalization (**Figure 2**; OR = 9.54 [8.14-10.94], FET p = 0.0184) whereas for those with C*04:01, it appears a white ethnic background and a more advanced age (≥65 years old) may be predisposing to ICU hospitalization (**Figure 2**; OR = 14.25 [12.24-16.26], FET p = 0.0053 and OR = 9.66 [8.27-11.05], FET p = 0.0188, respectively).

**Figure 2.**
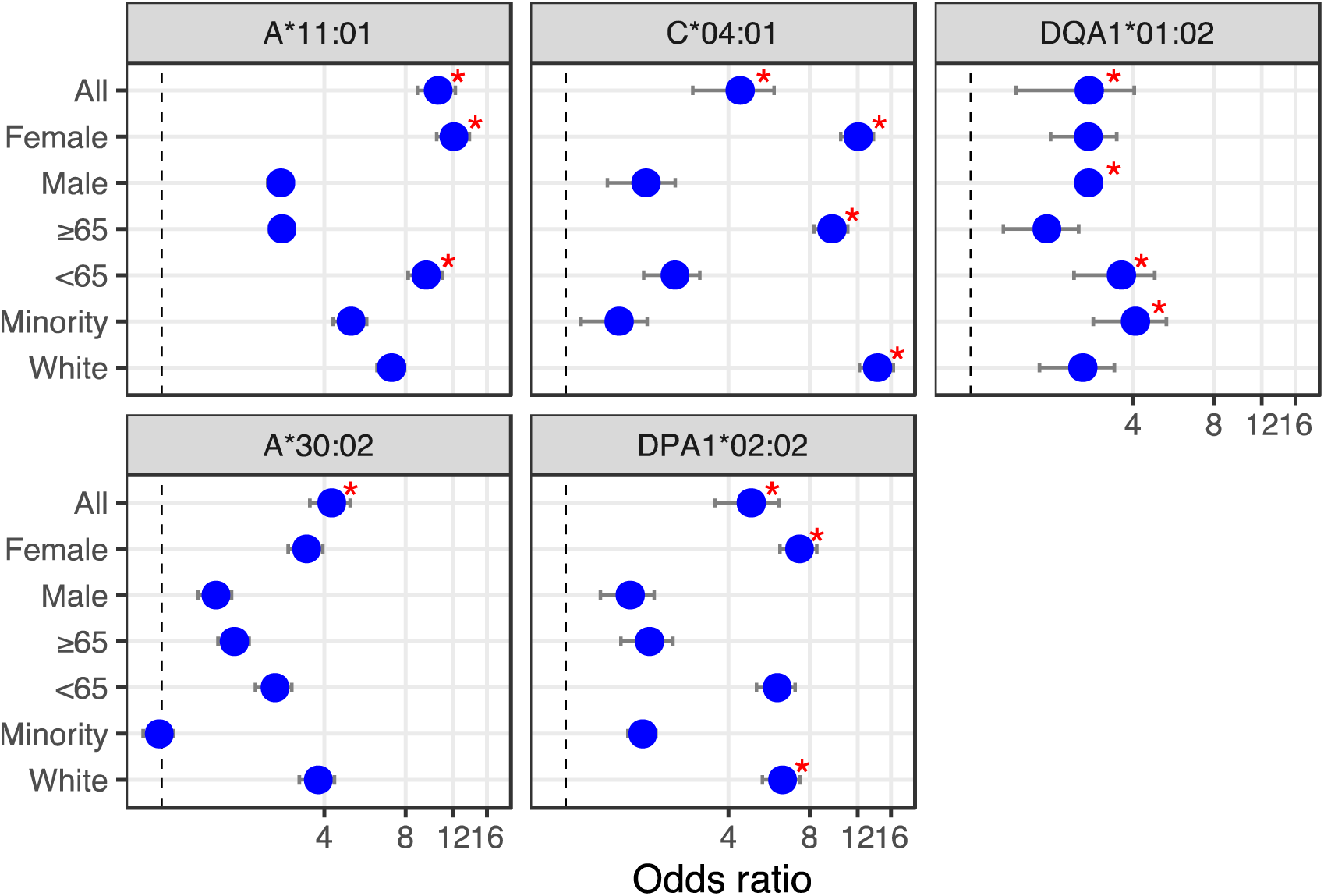
HLA alleles – demographics combinations with diagnostic (bottom) or prognostic (top) potential in a COVID-19 cohort. We calculated the odds ratio (OR) for each HLA-I and HLA-II alleles observed in 10% or more of patients, and plotted OR and the influence of demographics for HLA alleles showing significant associations (from **Tables 1 and 2**). First, looking at the influence of demographic characteristics such as sex (male/female), age (65 years old or above/less than 65 years old) and ethnicity (minority/white ethnic background) on the susceptibility of patients with these alleles to test positive for COVID-19 (lower two panels), and on the risk associated with ICU hospitalization (upper three panels). Red asterisks indicate significant demographic characteristics (Fisher’s Exact Test) not corrected for multiple hypothesis tests.

## DISCUSSION

We have previously identified the DPA1*02:02 class II allele as being prevalent in two other and independent cohorts, with patients of undisclosed ethnic background, but hospitalized in Wuhan, China [19]. Of populations with reported allele frequencies and an appreciable sampling size (≥100 individuals), only Hong Kong Chinese and Japanese have DPA1*02:02 allele frequencies (55.8% and 43.5%, respectively; [25,26]) above its observed frequency (29.0%) in the COVID-19 positive cohort analyzed herein. The frequency of this allele in other qualifying populations tends to be generally lower, including in South African (Worcester, 15.6%), Norwegians (14.0%), Mexico Chiapas Lacandon Mayans (6.7%), United Kingdom Europeans (4.3%) and Spain Navarre Basques (2.2%). We note that the ethnic background of the Overmyer *et al* [20] cohort is heterogeneous, and white individuals (of unknown ancestry) represent 45.1% and (11/29) 37.9% of the COVID-19 positive cohort and its DPA1*02:02 subset, respectively. In contrast, Asians represent only a minority of the cohort (1.9%) and DPA1*02:02 subset (1/29∼3.4%). It is important to note that, in the Overmyer cohort, DPA1*02:02 is not statistically associated with increased risk of hospitalization. The significant enrichment of this allele in the COVID-19 positive vs. negative cohorts (FET p= 0.0174) across all individuals, but also when looking only at females (FET p= 0.0326) or white individuals (FET p= 0.0491), and not any other demographics, may prove an important disease marker, which would need to be validated with additional datasets and in independent studies.

There are reports of disease associations with DPA1*02:02, DQA1*01:02, C*04:01 and A*11:01, but they are few. Of note, the association of all aforementioned alleles with narcolepsy [10,11] and a known trigger for this auto-immune disease includes upper-airway infections and influenza vaccinations [27-32]. Susceptibility to narcolepsy may in fact be an indirect effect of HLA class I and the HLA class II DP isotype in response to viral and bacterial infections, including from influenza and *streptococcus* [10,27,33,34]. It has since been reported that HLA-A*11 may be a susceptibility allele to influenza A(H1N1)pdm09 infection in some populations [35] while another report implicates HLA-I allele C*04:01 with high HIV viral loads [36]. Further, it was recently demonstrated that MHC class II DR, DQ and DP isotypes play a role in mediating the cross-species entry of bat influenza viruses *in vitro* in human/animal cell lines and in mice where engineered MHC-II deficiency made them resistant to upper-respiratory tract infections [12]. It is therefore not a stretch to envision an involvement from these HLA class II isotypes in controlling the cellular entry of a broader range of viral agents *in vivo*.

In a recent study examining HLA susceptibility based on SARS-CoV-2 derived peptide (epitope) binding strengths [37], the HLA-I allele A*11:01 was *in silico* predicted to bind a large number of SARS-CoV-2 derived peptides (n=750) with varying affinity [IC50 range 4.95 – 498.19, median = 149.62, mean = 182.28], and has been experimentally validated to bind SARS-CoV-2 peptide GLMWLSYFV (Tables S4 and S7 in Nguyen *et al*. [37]). In contrast, C*04:01 was only predicted *in silico* to bind six SARS-CoV-2 peptides and at higher IC50 ranges [167.65 – 469.30, median = 291.06, mean = 299.01] (Table S7 in Nguyen *et al*. [37]) suggesting a more limited ability to present epitopes to T cells and mount an appropriate immune response.

There have been a number of reports published on HLA alleles − COVID-19 associations this past year, and on cohorts from many jurisdictions including China [18], Italy [13,14,16] and the UK [17]. Wang *et al*. [18] compared the HLA allele frequencies between a cohort of 82 Chinese individuals and a control population of bone marrow donors previously studied by the same group. Novelli and co-workers [14] HLA typed a cohort of 99 Italian COVID-19 patients, and associated the observed allele frequencies with the HLA types in a reference group of 1,017 Italian individuals also previously studied by the same group. Correale *et al*. [13] and Pisanti *et al*. [16] followed a different strategy; these two independent studies leveraged population scale genomics data retrieved from the Italian Bone-Marrow Donors Registry and the National Civil Protection Department. They correlated background HLA allele frequency data with mortality and morbidity rates across Italy to reach at starkly different conclusions on which HLA alleles may play a role in disease etiology and progression. Disagreement between these two studies (also distinct from the results of the other Italian study by Novelli *et al*.) highlight the importance of large cohorts with matched samples to infer the patient HLA alleles with better statistical significance. Poulton *et al*. [17] characterized the HLA types of 80 COVID-19 patients in the UK on waiting lists for transplantation, and compared observed allele frequencies in comparison to a cohort of 10,000 deceased organ donors and a separate cohort of 308 SARS-CoV-2-negative individuals also on waiting lists for transplantation, the latter representing a matched demographics for the COVID-19 patients in their cohort. Interestingly, this is the only study that had any overlap between the alleles they flagged as being statistically significant and the lists published by other studies cited above. Not surprisingly, the alleles they listed do not intersect with the alleles identified and presented herein and the three alleles we published earlier on a very small group of only eight patients. It is nonetheless intriguing to find little to no HLA allele overlap between these reports, including with those associated with the 2003 SARS outbreak, a related respiratory disease caused by coronavirus [15,38,39]. This could be explained, at least partially, by geographical differences and varying population allele frequencies in those cohorts, relatively small cohort sizes (<100 patients), differences in experimentation setup and/or other factors, including comorbidity status, that may be acting independently of HLA.

## CONCLUSIONS

Here, we predict HLA-I and HLA-II alleles from publicly available COVID-19 patient blood RNA-seq samples and identified several putative biomarkers. In a previous study, we had observed one of these biomarkers (DPA1*02:02), and we postulate that patients with the allele may have an increased susceptibility for COVID-19. Further, other alleles (A*11:01, C*04:01, DQA1*01:02) may be prognostic indicators of poor outcome. However, although it is well established that patient HLA profiles play a significant role in the onset and progression of infectious diseases in general, we caution against drawing overreaching conclusions from regional, and often limited, observations. We note that recently published studies associating HLA alleles and COVID-19, by and large, disagree in their findings. We expect future studies with larger cohort sizes will help bring a clearer picture of the role of patient HLA profiles, if any, in COVID-19 susceptibility and disease outcomes.

## Data Availability

The RNA-seq datasets analysed during the current study are available in the ENA repository https://www.ebi.ac.uk/ena/browser/view/PRJNA660067 accessions: SRX9033799- SRX9033924. The associated clinical data are available in the GEO repository https://www.ncbi.nlm.nih.gov/geo accession: GSE157103

https://www.ncbi.nlm.nih.gov/geo/query/acc.cgi?acc=GSE157103

## LIST OF ABBREVIATIONS

FET: Fisher’s Exact Test
HFD-45: hospital-free days post 45 day followup (days)
HLA: human leukocyte antigen
ICU: Intensive Care Unit
KM: Kaplan-Meier

## DECLARATIONS

### Ethics approval and consent to participate

Not applicable

### Consent for publication

Not applicable

### Availability of data and materials

The RNA-seq datasets analysed during the current study are available in the ENA repository https://www.ebi.ac.uk/ena/browser/view/PRJNA660067 accessions: SRX9033799-SRX9033924. The associated clinical data are available in the GEO repository https://www.ncbi.nlm.nih.gov/geo: accession GSE157103

### Authors’ contributions

RLW designed the study, analyzed the data and wrote the manuscript. IB participated in the development of the study and co-wrote the manuscript. All authors have read and approved the manuscript.

## Acknowledgements

Not applicable

